# At the Edge: Disability, Crisis, and Non-Suicidal Self-Injury in Higher Education

**DOI:** 10.1101/2025.07.29.25332248

**Authors:** Geethika Kodukula

## Abstract

**Objective:** To examine the differences in the prevalence of Non-suicidal Self-Injury (NSSI) over the past year between Students with Disabilities (SWD) and their peers.

**Participants:** 128,310 students aged 18-35 years from universities throughout the United States.

**Methods:** Secondary data analysis obtained from the Healthy Minds survey (2020-2021) was analyzed using multiple logistic regression to estimate the odds of reporting NSSI among SWD and their peers. Covariates included: demographics, stressful life experiences (SLE), physical and mental health, substance use, and institutional characteristics.

**Results:** NSSI was significantly associated with disability; unadjusted odds ratio = 1.95 (1.8, 2.1) and adjusted odds ratio = 1.26 (1.2, 1.4). Being 2SLGBTQIA+, a history of physical abuse, loneliness, depression, and suicidal ideation increased the odds of NSSI 50% or more.

**Conclusions:** SWD were more likely to report NSSI in the past year compared to their peers. The results emphasize the need for holistic, targeted support for SWD.

## Introduction

Mental well-being can impact a college student’s personal health, academic success, relationships, and subsequently, their earning potential and quality of life (Johnson et al., 2018). Over the last two decades, the rate of diagnosis of mental health conditions among college students has been trending upward (Duffy et al., 2019), with symptoms of anxiety and depression doubling in 2021 compared to 2013 (Lipson et al., 2022). In addition to navigating the higher education landscape, when the COVID-19 pandemic hit, college students around the world, and particularly some subgroups, faced additional, unexpected, and complex layers of stress in 2020 (Centers for Disease Control and Prevention, 2023; C. Son et al., 2020). Severe psychological distress saw a sharp rise in the nascent months of the COVID-19 pandemic (Lee et al., 2020). This was also reflected in some coping mechanisms; studies found that risky alcohol drinking and alcohol use disorders spiked among students, compared to pre-pandemic levels (Kim et al., 2022; Lechner et al., 2021). In particular, some groups reported higher levels of isolation and worsened discrimination, such as international students, race and gender minorities and students with disabilities (Coduti et al., 2016; Lai et al., 2020; Rung et al., 2024; Trusty, 2023).

There are approximately 4.3 million undergraduate and 300,000 post-baccalaureate students with disabilities (hereafter, SWD) in the US (National Center for Education Statistics, 2023; Office of Disability Employment Policy). Although accessibility improved over the years and awareness campaigns combated discrimination, living in a disabled body in a society built around the needs of able-bodied people can lead to higher levels of life stress, lower life satisfaction, and other poor life outcomes (Lund, 2021). To wit, SWD report higher levels of food insecurity, childhood abuse, and intimate partner violence (Findley et al., 2016; Hughes et al., 2011; E. Son et al., 2020; Stott & Morrell, 2021). During the COVID-19 pandemic, SWD reported higher levels of psychosocial stressors such as anxiety and loneliness (Lund, 2020; McMaughan et al., 2021; Soria & Coca, 2023), and although some SWD benefited from the shift to online learning, existing digital divides widened during the pandemic (Butler & Savalli, 2021; Scott, 2020). Pre- and post-pandemic, the most commonly reported manifestations of stress among students with disabilities were frustration and loneliness, anxiety, depression, non-suicidal self-injury (hereafter, NSSI), and suicidal thoughts and behaviors (Aguilar & Lipson, 2021; Fleming et al., 2018).

NSSI is a serious public health problem that caused approximately 180,397 ER visits in the US in 2022 (National Center for Injury Prevention and Control, 2024). It is defined as ‘an act of deliberate harm to one’s own body tissue without the intent to die’ (Nock et al., 2006). Prevalence estimates range around 13%-30% in the student population (DeAngelis, 2015; Hamza et al., 2021; Klonsky, 2011; Swannell et al., 2014), but this is likely an undercount since NSSI is by nature a transient and private phenomenon, unpredictable in occurrence (Nock, 2010). Additionally, variations in definitions and descriptors across studies and information biases introduced by using large-scale self-reports or observations in a clinical setting make it difficult to establish the true prevalence.

Prevention of NSSI is an important part of suicide prevention, as it is both theoretically and empirically related to an increased risk of suicidal behavior (Joiner, 2005; Mars et al., 2019; Turner et al., 2013; Van Orden et al., 2010). However, the motivations behind each behavior are distinct; the most commonly reported motive for NSSI is alleviation of an extremely negative state (Klonsky, 2011; Nock, 2010). However, it is not without important consequences such as an increased risk of substance use, eating disorders, poorer academic achievement, and subsequently lower economic potential (Auger et al., 2022; Kiekens et al., 2018).

Research on NSSI among young adults is important to prevent accidental mortality, reduce morbidity, and understand antecedent behaviors that are modifiable. Understanding NSSI among SWD is particularly important because they are at the intersection of two vulnerable groups but are often overlooked and under researched. They also encounter higher rates of experiences that are associated with NSSI and STB, such as childhood abuse, intimate partner violence, higher levels of mental distress, depression, and substance use disorders (Casseus et al., 2021; Coduti et al., 2016; Scherer et al., 2016).

The current study utilizes a dual framework to understand the mental health of SWD during the pandemic: the acute on chronic stress framework and the minority stress model (Gabrielli et al., 2013; Meyer, 2003). We hypothesize that although the COVID-19 pandemic was a highly impactful acute stressor on all students, those living with the chronic stress of being a minority like SWD, experienced compounded effects. Figure i visually represents the conceptual model integrating acute-on-chronic stress and minority stress frameworks.

**Figure i.**
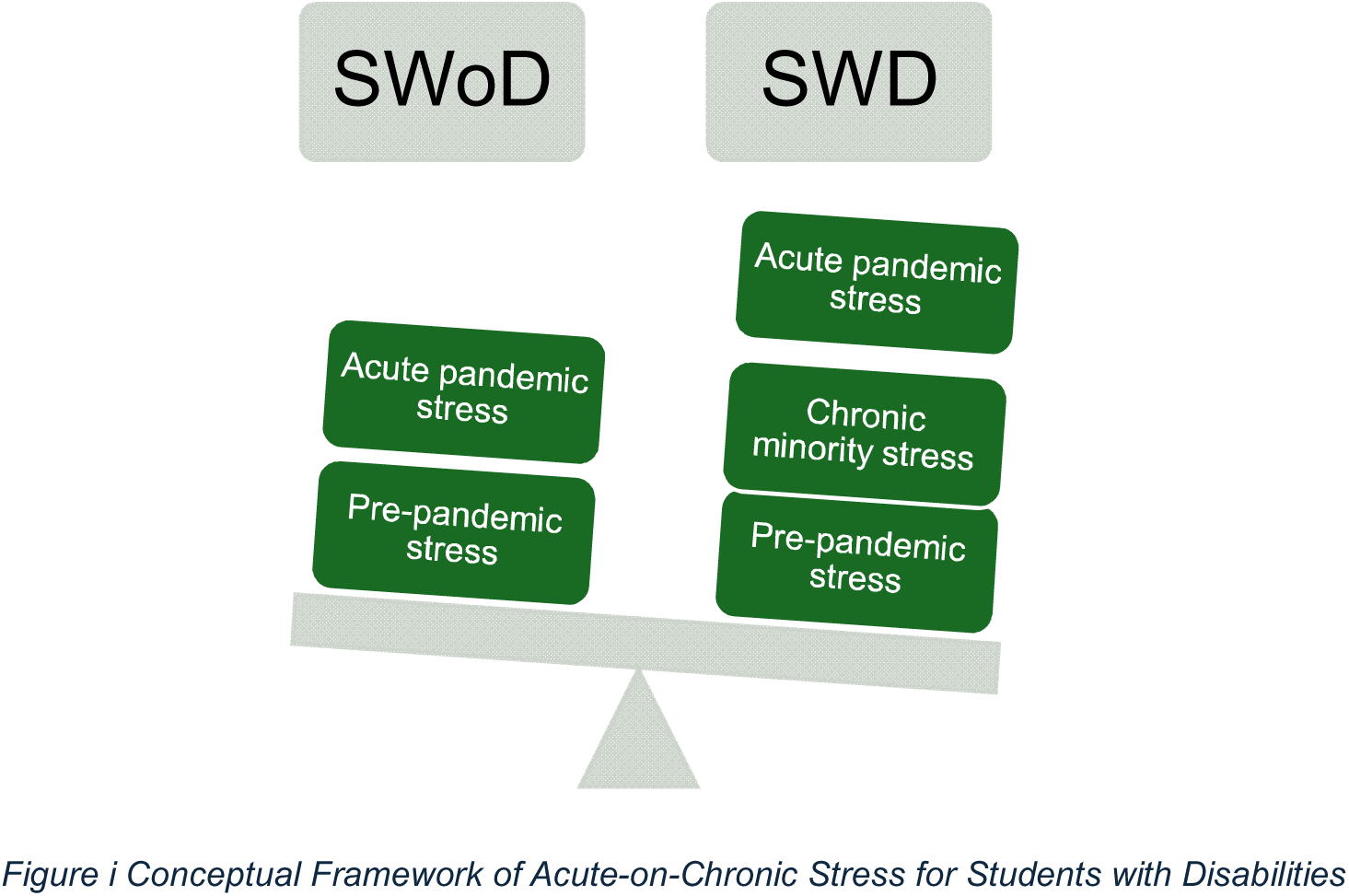
Conceptual Framework of Acute-on-Chronic Stress for Students with Disabilities

## Materials and Methods

### Research Design

This study used secondary data analysis to answer the research question: during the COVID-19 pandemic, did students’ self-reported NSSI in the previous year differ by their disability status?

The data used in this study were obtained from the national survey by the Healthy Minds Network (HMN) (Healthy Minds Network, 2021). Among institutions that sign up, the survey is sent to students at random. Nonresponse weights were calculated by the HMN based on institutional administrative data, with each school in the national sample given equal aggregate weight in the national estimates (Eisenberg et al., 2020).

#### Institutional Review Board Approval

Since the analysis uses secondary data, an exempt protocol was approved by the Institutional Review Board at [institution redacted for review] (IRB: #XXX). In the recruitment emails sent by the HMS, students are provided with information regarding the minimal risk they may experience in responding to sensitive questions and are required to provide express consent to continue with the survey. For those who did not consent, the survey process terminated with information provided for local counseling services.

### Sample

Participants included college students from across the US who responded to the Fall 2020 or Winter 2021 surveys. Although “emerging adulthood” is defined as those who are aged 18-25 years old (Arnett, 2000), we extended the inclusion range to 35 because SWD often enter higher education later than their peers and sometimes take longer to complete their degrees (Newman et al., 2011; Wessel et al., 2009). With 128,310 total observations, including 9,409 SWD and 27,984 reporting NSSI, the sample size substantially exceeded what would be required to detect even small effects.

### Measures

Independent variable *disability* was ascertained using the question: *Are you registered with the office for disability services on this campus as having a documented and diagnosed disability?*

Dependent variable, *NSSI*: A binary outcome was constructed using responses to the question: *In the past year, have you ever done any of the following intentionally? (This question asks about ways you may have hurt yourself on purpose, without intending to kill yourself*). Nine common NSSI methods were listed^1^, respondents could select one or more options, *other*, or *none*.

Variables to be included in the primary analysis as covariates were selected based on extant literature, drawn from the three core modules of the Healthy Minds Study.

1) Demographics: age, sex, gender, sexual or romantic orientation, race-ethnicity, and international student status.
2) Stressful Life Experiences (SLE): having a diagnosed chronic illness^2^, food insecurity in the past year (Hager et al., 2010), lifetime exposure to physical abuse, any experiences of assault or violence in the previous year (physical, mental, or sexual), a change in financial situation due to the COVID-19 pandemic, and whether poor mental health impacted their academics at least one day in the past month.
3) Mental health measures: Validated instruments like the PHQ-9 (Kroenke et al., 2001), GAD-7 (Spitzer et al., 2006), the revised UCLA loneliness scale (Hughes et al., 2004), and the SCOFF disordered eating scale (Morgan et al., 2000) were utilized to establish the general and recent mental health of the respondents. All scales showed moderate to high internal consistency in this sample, with Cronbach’s alphas ranging from 0.8 to 0.92, except for the SCOFF scale (0.56). But due to the high comorbidity between disordered eating NSSI, it was included in the analysis (Cornell University College of Human Ecology, n.d.). Additionally, one of the three questions from the National Comorbidity Survey Replication (Kessler et al., 2005) was used to measure suicidal ideation in the past year.
4) Self-reported measures of alcohol use, cigarette smoking, and drug use^3^.
5) Two institutional variables were included to account for geographic variations and related lockdown policies at the school level, as well as correlations with resource availability: geography and sector (private/public).

### Statistical Analysis

All statistical analyses were conducted using SAS 9.4 software™. Of the 128,310 observations, 12.7% were missing NSSI data and 7.4% were missing disability status. Missing data patterns in key variables were assessed by creating indicators for non-response, then tested in bivariate analyses against demographic variables. By default, the regression models used complete case analysis, resulting in an confidence level.

## Results

Table 1 presents sample characteristics overall and between the groups of students who reported any non-suicidal self-injury in the past year. Approximately 8.5% (7.8, 9.2) of the sample reported having a registered disability and one in four students in the sample reported a 12-month prevalence of non-suicidal self-injury (23.7, 25.8). Since the bivariate analysis of indicator variables for missing disability information showed that the data was not missing completely at random (MCAR), a complete case analysis was used to conduct the multiple logistic regression.

**Table 1.**
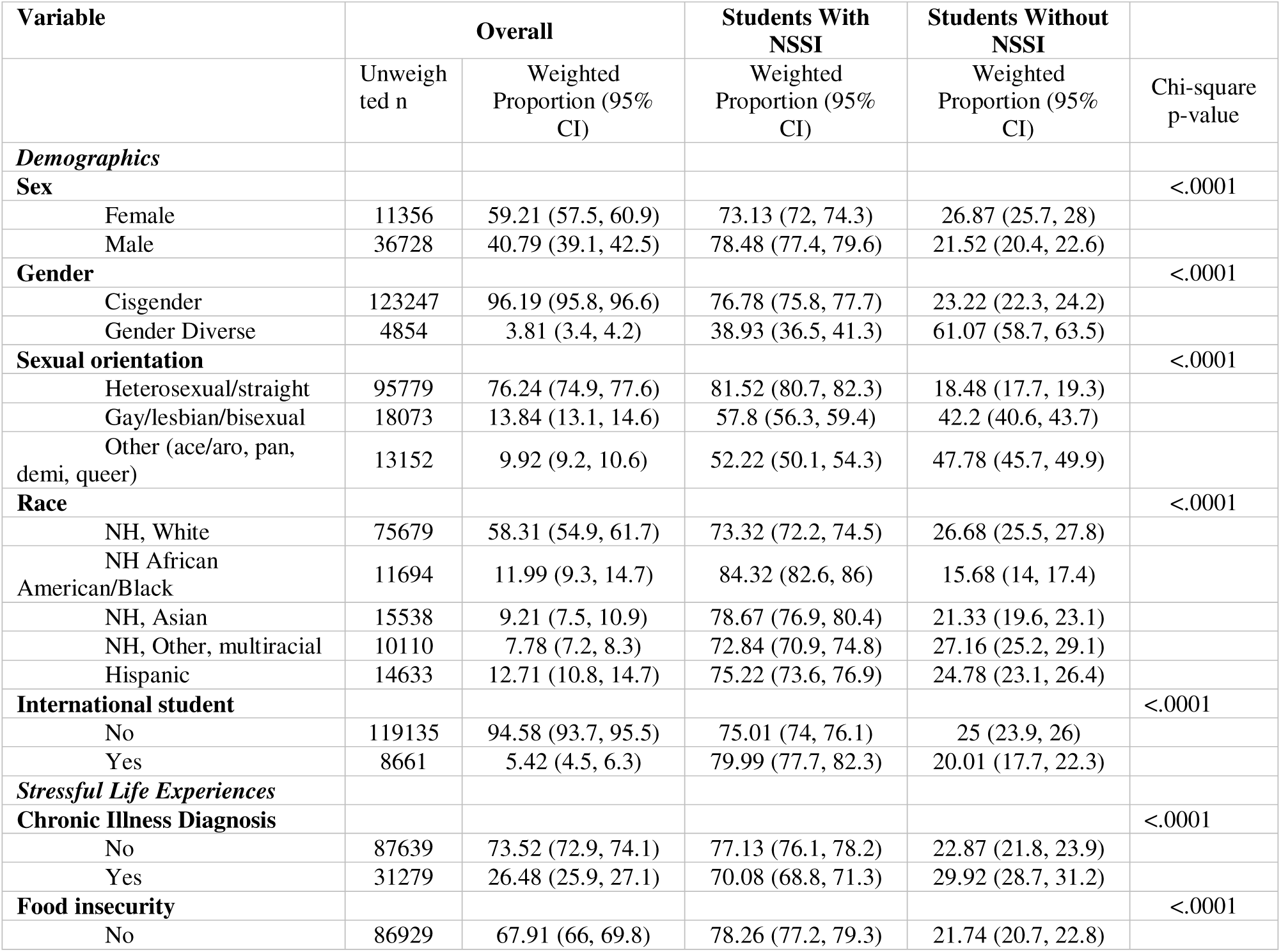
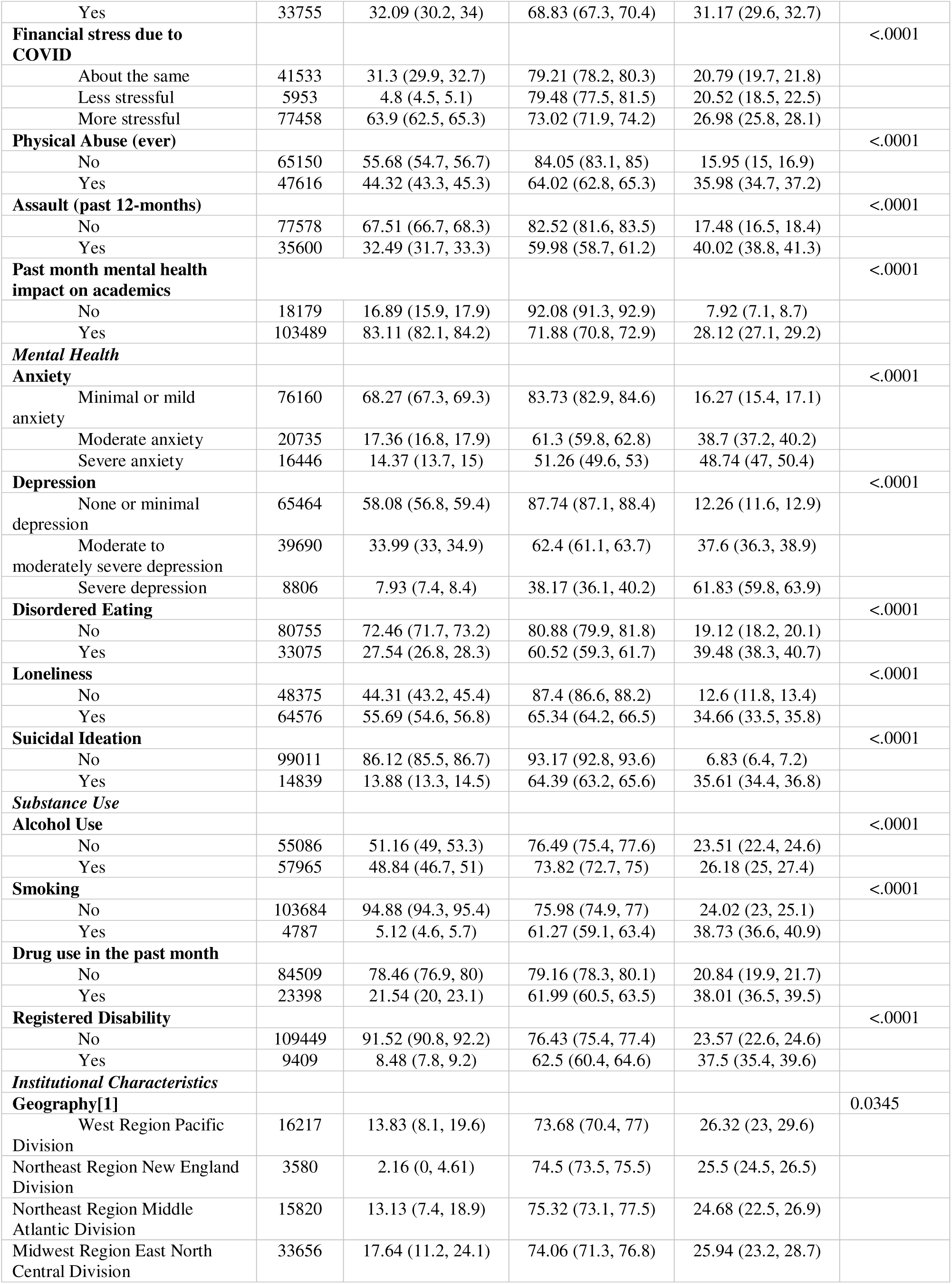
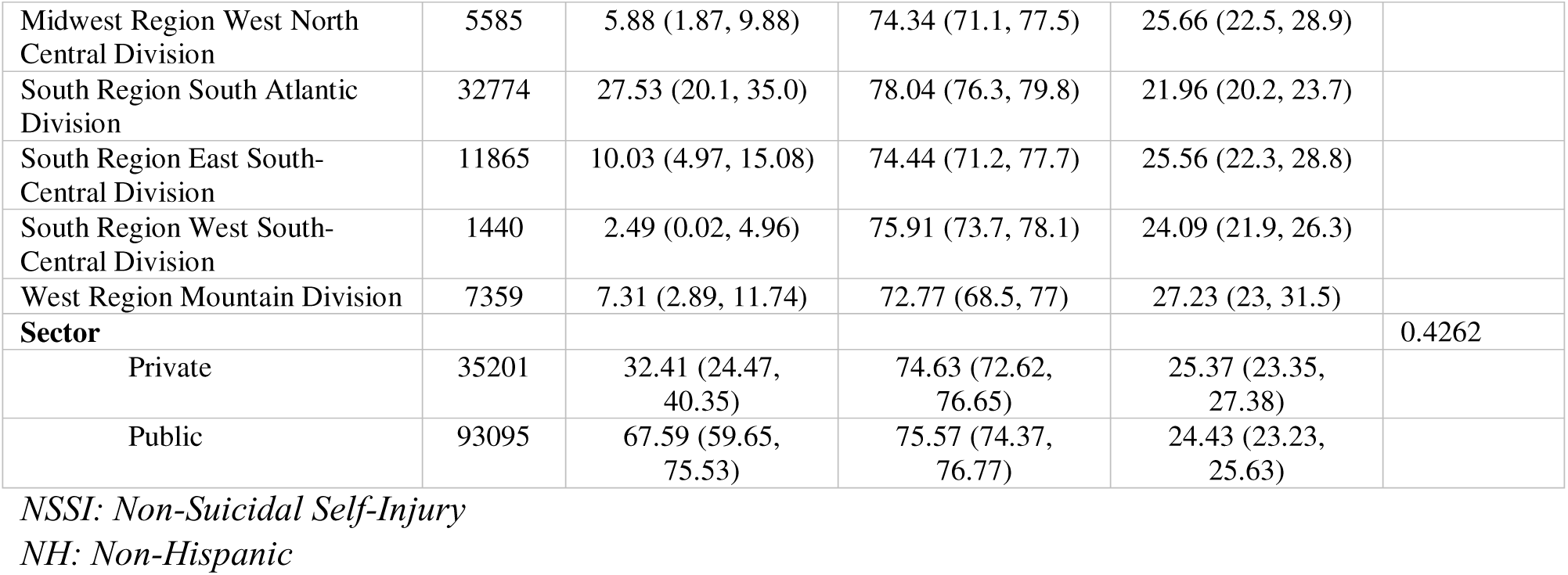
Sample Characteristics by Non-Suicidal Self-Injury (n =128,310)

### Demographics

The weighted average age for this age-delimited sample was 21.9 years (21.7-22.1). The sample was 60% female, 96% cisgender, 76% heterosexual, and 58% non-Hispanic White. A cross-tabulation of variables by a history of NSSI showed predictable trends: a higher proportion of female students endorsed having engaged in NSSI in the previous year compared to males (26.9% vs 21.5%).

Approximately 23.2% of cisgender students reported NSSI while the proportion was higher among transgender/non-binary/queer students (61.1%). Similarly, when broken down by sexual or romantic orientations, 42.2% of LGB students, and 47.8% of other sexual/romantic orientations (queer, aromantic, asexual, etc.) reported a history of NSSI compared to 18.5% of heterosexual students.

The following variables were found to have a statistically significant association with NSSI: age, sex assigned at birth, race, and sexual orientation, chronic illness, financial stress, food insecurity, mental health screening scores: depression, anxiety, loneliness, disordered eating and substance use: alcohol, tobacco, and illicit drug use and disability status (Table 1). For the multiple regression, the continuous variable age was standardized, the reference value for all binary outcomes was ‘No’ and among categorical variables with more than two values, the group with the largest proportion was selected as the reference group. Results of the regression analysis are presented in table 2.

**Table 2.** Results from Multiple Logistic Regression to Predict Non-Suicidal Self-Injury among College Students (n = 128,310)

| Effect | Adjusted Odds Ratio [95% Confidence Interval] |
| --- | --- |
| Demographics |  |
| Age | 0.99 [0.99, 0.99]** |
| Sex (Female) |  |
| Male | 1.01 [0.95, 1.08] |
| Gender (*Cisgender*) |  |
| Gender Diverse | 1.79 [1.57, 2.03] ** |
| Sexual Orientation (Heterosexual) |  |
| Gay/Lesbian/Bisexual | 1.65 [1.53, 1.78] ** |
| Other | 2.13 [1.96, 2.31] ** |
| Race (*NH White*) |  |
| NH African American/Black | 0.54 [0.48, 0.61] ** |
| NH, Asian | 0.88 [0.78, 0.99] * |
| NH, Other, Multiracial | 0.83 [0.75, 0.92] * |
| Hispanic | 0.83 [0.76, 0.9] ** |
| International Student (*Domestic Student*) | 1.29 [1.1, 1.51] * |
| Stressful Life Experiences |  |
| Chronic Illness Diagnoses (*No*) | 1.12 [1.06, 1.18] * |
| Food Insecurity (*No*) | 0.94 [0.88, 1.01] |
| Financial Stress Due To Covid (*No Change*) |  |
| Less Stressful | 1.01 [0.86, 1.18] |
| More Stressful | 0.88 [0.84, 0.93] ** |
| Physical Abuse Ever (*No*) | 1.66 [1.57, 1.76] ** |
| Assault In the Past Year (*No*) | 1.49 [1.41, 1.58] ** |
| Past Month Mental Health Impact on Academics (*No*) | 1.63 [1.45, 1.83] ** |
| Mental Health |  |
| Anxiety (None Or Minimal) |  |
| Moderate Anxiety | 1.37 [1.29, 1.46] ** |
| Severe Anxiety | 1.37 [1.27, 1.48] ** |
| Depression (*None or Minimal*) |  |
| Moderate | 1.78 [1.66, 1.91] ** |
| Severe | 2.5 [2.22, 2.82] * |
| Disordered Eating (*No*) | 1.41 [1.34, 1.48] ** |
| Loneliness (*No*) | 1.54 [1.45, 1.64] ** |
| Any Suicidal Ideation (*No*) | 3.27 [3.01, 3.56] ** |
| Substance Use |  |
| Alcohol Use (*No*) | 0.97 [0.92, 1.03] |
| Smoking (*No*) | 1.2 [1.06, 1.36] * |
| Drug Use (*No*) | 1.44 [1.36, 1.54] ** |
| Institutional Characteristics |  |
| Geography (*Western Region Pacific Division*) |  |
| Northeast, New England Division | 0.99 [0.81, 1.2] |
| Northeast, Middle Atlantic Division | 0.87 [0.69, 1.1] |
| Midwest, East North Central Division | 0.94 [0.77, 1.14] |
| Midwest, West North Central Division | 1.01 [0.85, 1.21] |
| South, South Atlantic Division | 0.97 [0.78, 1.19] |
| South, East South-Central Division | 0.89 [0.74, 1.07] |
| South, West South-Central Division | 0.93 [0.77, 1.13] |
| West, Mountain Division | 0.94 [0.78, 1.13] |
| **Disability (No)** | **1.26 [1.15, 1.4] \*\*** |

Reference category in parentheses
NH: Non-Hispanic
\* *p*-value <.05
\*\* *p*-value <.0001

Bivariate logistic regression showed that reported disability had an unadjusted odds ratio of 1.95 [1.8-2.1]. The multiple regression predicting any NSSI in the previous year based on disability status while controlling for demographics, SLEs, mental health, substance use, and institutional variables showed that students with a registered disability were 25% more likely to have had a past-year NSSI compared to those without a disability. Therefore, the hypothesis that students with disabilities did not significantly differ in terms of their past-year NSSI was rejected.

Most of the variables that had a significant effect on NSSI in the bivariate analysis continued to do so in the multiple regression, but with some modified effect sizes. Sex was no longer a statistically significant predictor of NSSI, but gender-diversity and minority sexual or romantic orientations both increased odds compared to their respective peers (1.79 [1.57, 2.03] and 1.65 [1.5, 1.8] respectively).

Belonging to a race other than non-Hispanic White decreased the odds of reporting NSSI, and the odds varied between levels of the race variable (AOR range: [0.53, 0.89]).

Notably, students who screened positive for disordered eating were 40% more likely to report NSSI, and the effect severe depression had on odds of having engaged in NSSI was much higher (AOR = 2.4 [2.1, 2.7]) than it was for severe anxiety (AOR = 1.36 [1.3, 1.5]). Students who reported that their mental health impacted their academics at all had 64% [1.5, 1.8] increased odds of reporting NSSI compared to those reported that there was no impact. As can be expected, any exposure to violence whether lifetime or recent increased one’s odds of reporting NSSI: (AOR range: [1.5, 1.7]). Drug use showed a stronger association with NSSI (AOR = 1.45 [1.4, 1.6]) compared to smoking (AOR = 1.2 [1.1, 1.4]). The variable that had the largest effect was suicidal ideation (AOR = 3.3 [3.0-3.5]).

The model had moderate predictive power with a Nagelkerke’s R^2^ of 0.6735 (Nagelkerke, 1991). Two potential interaction effects, between disability and race, and between disability and reporting that mental health impacted one’s academics were tested and found not statistically significant.

## Discussion

This study illuminates that despite accounting for most things that impact one’s mental health, having a disability still meant increased odds of reporting past-year self-injury. Consistent with Coduti et al. (2016), the study found that SWD reported NSSI at higher proportions than their non-disabled peers. By using a national weighted dataset, our study examined this relationship particularly during a crisis, building upon Dr. Lund’s proposal to use minority stress theory for explaining suicidality (and similar behaviors) among PWD (Lund, 2021). It also operationalizes a dual framework with acute-on-chronic stress and minority stress mechanisms (Gabrielli & Lund, 2020; Lund, 2021b). While one other study, Halpern-Manners (2022) noted declines in physical and mental health among students with disabilities during the pandemic, they did not focus on NSSI, making this a novel contribution.

Among the considered demographics, belonging to a gender or sexual orientation minority group was found to increase odds of NSSI even after adjusting for correlates. These results align with existing research on minority stress and address the compounded stress that SWD experienced during the COVID-19 pandemic. These findings also align with Hatzenbuehler’s psychological mediation framework (2009) which identifies how stigma-related stress can lead to emotional dysregulation, one of the most commonly cited reasons for engaging in NSSI (Nock & Favazza, 2009). Based on the intersectional framework (Crenshaw, 1989), it stands to reason that SWD who are LGBTQ+ and/or ethnic minorities may face compounded stressors. One notable exception to the trend that has been affirmed by seminal researchers like Klonsky & Muehlenkamp (2007) was the finding that non-Hispanic White students were more likely to have engaged in NSSI compared to their peers. This suggests that some community-level protective factors for race and ethnic minorities may be acting as protective factors, highlighting the continued need for clear race-ethnicity distinctions in public health research.

Among the SLEs examined, one interesting finding was regarding the change financial stress due to COVID-19 pandemic. Students who reported that their financial situation was somewhat or much more stressful due to COVID-19 were *less* likely to report past year NSSI compared to those who said their situation remained the same. While factors like increased or newfound access to familial or governmental support could have played a part, further studies that are longitudinal and/or qualitative in nature can help understand this mechanism empirically.

One unexpected result was that alcohol use was not statistically significant as a predictor in this sample, contrary to current opinions (Jacobucci & Ammerman, 2023; Shepherd et al., 2023). This may be due to the high drinking rates in the sample (48.84%) affecting the ability to detect meaningful effects.

Lechner et al. (2021) found that drinking among college students spiked compared to pre-pandemic levels. Another possible reason is the difference in temporality (past-year NSSI vs drinking in the previous fortnight). On the other hand, drug use and smoking increased odds of reporting NSSI, similar to findings by Serras et al. (2010), Goñi-Sarriés et al. (2025).

### Limitations

These findings must be considered in the context of a few limitations.

- All measures were self-reported, prone to social desirability and recall biases.
- Data were not missing completely at random (MCAR), with nearly 23% of the original dataset excluded from the regression model due to missing data on key variables and the study used complete case analysis (CCA). If the missingness is meaningfully related to unobserved variables, there is loss of information and potential for bias.
- Similarly, using a student’s registered disability as a proxy for having a disability is less than perfect because it underestimates the prevalence and effect of disability; many students with disabilities do not register with the campus accessibility services, though this varies by type of disability and is due to a myriad of reasons like not being aware of services, stigma, etc. (Newman & Madaus, 2015).
- Temporality: the question regarding NSSI spans the previous year, which could introduce some bias if it occurred pre-pandemic.

### Strengths

The limitations are balanced by the strengths of this study:

- To our knowledge, it is the first of its kind examining severe psychological distress and non-suicidal self-injury in a under-researched population who are at higher risk of distress than average. It is a telling snapshot of the state of mind of young adults in college in the face of a seismic change.
- Notably, the theoretical and conceptual frameworks proposed were validated, since SWD not only showed higher levels of proximal stressors (depression and anxiety levels in the previous two weeks, food insecurity in the past year), but also higher levels of chronic stressors (lifetime abuse, instances of victimization).
- The use of validated psychometric instruments and a national dataset while accounting for non-response and complex survey sampling lends methodological rigor and improves the generalizability of the results. Comprehensive covariate adjustment spanning demographic, intrapersonal, interpersonal, and institutional variables helps identify the independent effect of disability on NSSI.

### Public Health Implications

The natural next step is to identify ways that can mitigate this risk. A collaborative effort from different units in a university can help support SWD in the following ways:

For campus accommodation services: 1) Training staff in identifying warning signs of NSSI and those that may be specific to the SWD experience. 2) Coordinating peer-support mental health groups that can introduce students to connect through shared experience and support those at different risk levels than themselves. 3) Targeted screening protocols that are informed by risk factors unique to SWD such as those identified in this study, for example, those who are presently food insecure.
For campus health centers: 1) Integrating screening for NSSI into routine health visits processes with attention to SWD, who often use the centers more often than their peers. 2) Integrating the student accessibility/accommodation services on campus with mental health services, and 3) Implementing a trauma-informed approach to all healthcare, cognizant of the higher rates of financial stress, abuse, and assault that SWD experience. 4) Screening for co-occurring substance use and NSSI to identify those at higher risk of harm.
For administrators: 1) Developing emergency protocols to use during future crisis using disability community based participatory research (CBPR). 2) Allocate resources to maintain support services to SWD during times of disruption, such as telehealth services. 3) Campus-wide awareness programs addressing stigma around disability and NSSI individually and targeted workshops for SWD to improve protective factors such as resilience and belonging.
For surveillance systems: data disaggregated by disability type, level of functional impact, and experiences of discrimination or exclusion based on disability should be collected to illuminate the differences between the student groups better and to address those gaps more effectively instead of a one-size-fits-all approach to research and analysis.

Health and academic equity require equal opportunity and equal access to resources. When any disaster strikes, the first and most severely affected are the marginalized people in our communities.

Students with disabilities face unique challenges in the world, and all efforts possible should be made to ensure their health and academic success, especially under unforeseen and large-scale circumstances. In addition to living with the minority stress of being a student with disabilities, acute stressors that change or displace care and accessibility for students with disabilities can have serious mental health implications. Some suggestions to help mitigate this problem are: i) including simple and confidential screening for NSSI and suicidal ideation during all primary health check-ups, especially at campus medical centers. ii) Targeted intervention and awareness programs with tips to manage mental health particularly geared towards students with disabilities. iii) Finally, academic institutions should shoulder more responsibility to make sure that students are not experiencing such levels of psychological distress by using regular needs assessments, keeping a well-staffed psychological clinic on campus, and training teachers and staff like residential assistants to recognize signs that a student maybe seriously thinking of suicide or self-injury. This study is a step towards bridging the knowledge gap about severe psychological distress among students with disabilities and highlighting the modifiable factors impacting all students and their potential academic success. These results point to multiple avenues for disability accommodations offices to include in intervention and awareness programs by collaborating with other groups on campus to reach the multiply marginalized students who are prone to experiencing compounded stress.

## Generative Artificial Intelligence

The application Claude 0.11.6 was used to check grammar and improve flow.

## Data availability statement

The data that support the findings of this study are available from the Healthy Minds Network but restrictions apply to the availability of these data, which were used under license for the current study. Data are not publicly available but may be obtained upon reasonable request and with permission from the Healthy Minds Network (https://healthymindsnetwork.org/research/data-for-researchers/).

## Footnotes

1 The options were: cut myself, burned myself, scratched myself, punched or banged myself, pulled my hair, interfered with wound healing, carved words or symbols into skin, rubbed sharp objects into skin, punched or banged an object to hurt myself.

2 Conditions listed: Diabetes, High blood pressure, Asthma, Thyroid disease, Gastrointestinal disease, Arthritis, Sickle cell anemia, Seizure disorders, Cancers, High cholesterol, HIV/AIDS, Other autoimmune disorders, Other chronic diseases

3 Marijuana, cocaine, heroin, opioid pain relievers, benzodiazepines, methamphetamines, other stimulants, MDMA, ketamine, LSD, psilocybin, kratom, or athletic performance enhancers. analytic sample of 99,034 observations for the final model. All analyses accounted for complex survey sampling using survey weights and clustered design. All estimate intervals presented are at 95%

